# Evaluation of “stratify and shield” as a policy option for ending the COVID-19 lockdown in the UK

**DOI:** 10.1101/2020.04.25.20079913

**Authors:** Paul M McKeigue, Helen M Colhoun

## Abstract

Although population-wide lockdowns have been successful in slowing the COVID-19 epidemic, there is a consensus among disease modellers that keeping the load on critical care services within manageable limits will require an adaptive social distancing strategy, alternating cycles of relaxation and reimposition until a vaccine is available. An alternative strategy that has been tentatively proposed is to shield the elderly and others at high risk of severe disease, while allowing immunity to build up in those at low risk until the entire population is protected. We examine the performance required from a classifier that uses information from medical records to assign risk status for a such a stratify-and-shield policy to be effective in limiting mortality when social distancing is relaxed.

We show that under plausible assumptions about the level of immunity required for population-level immunity, the proportion shielded is constrained to be no more than 15% of the population. Under varying assumptions about the infection fatality ratio (from 0.1% to 0.4%) and the performance of the classifier (3 to 4.5 bits of information for discrimination), we calculate the expected number of deaths in the unshielded group. We show that with likely values of the performance of a classifier that uses information from age, sex and medical records, at least 80% of those who would die if unshielded would be allocated to the high-risk shielded group comprising 15% of the population. Although the proportion of deaths that would be prevented by effective shielding does not vary much with the infection fatality ratio, the absolute number of deaths in the unshielded varies from less than 10,000 if the infection fatality rate is 0.1% to more than 50,000 if the infection fatality rate is as high as 0.4%.

For projecting the effect of an optimally applied stratify-and-shield policy, studies now under way should help to resolve key uncertainties: the extent to which infection confers immunity, the prevalence of immunity, the infection fatality ratio, and the performance of a classifier constructed using information from medical records. It is time to give serious consideration to a stratify-and-shield policy that could bring the COVID-19 epidemic to an end in a matter of months while restoring economic activity, avoiding overload of critical care services and limiting mortality.

## Background

The Sars-CoV-2 epidemic will be over only when the level of immunity in the population reaches a level where the effective reproduction number falls below 1 through natural immunity or vaccination. It is expected to be at least a year before an effective vaccine can be developed and tested adequately. On 23 March 2020 stringent social distancing measures (termed lockdown) were imposed in the UK so as to prevent the epidemic from overwhelming the National Health Service and to reduce immediate impact on deaths. Modelling studies for the Scientific Pandemic Influenza Group on Modelling (SPI-M) have suggested that an adaptive strategy “alternating between periods of more and less strict social distancing measures” may be needed to keep the number of critical care cases within capacity until the epidemic ends [1]. Concern about the economic and social costs of this intermittent lockdown strategy has motivated exploration of other approaches for managing the epidemic from this point forward.

One suggested strategy for exit from the lockdown has been to undertake weekly testing of the entire population for SARS-CoV-2 RNA, followed by isolation of those who test positive and their contacts until the epidemic is suppressed [2]. Another option that has been tentatively proposed is to shield those at high risk while allowing immunity to build up in those at low risk [3]. A a briefing dated 26 February from the Scientific Advisory Group on Emergencies (SAGE) alluded to this “stratify-and-shield” approach [4]:

> *An additional strategy would be to apply more intense measures on those age or risk groups at most risk of experiencing severe disease (e*.*g. household isolation of those over 65, special measures around care homes). The majority of the population would then develop immunity, hopefully preventing any second wave,while reducing pressure on the NHS. However, SPI-M-O has not looked at the likely feasibility or effectiveness of such methods*.

In subsequent modelling studies for SPI-M, interventions including less stringent “social distancing” of those over 70 years of age were assessed based on assuming modest reductions in contacts with people outside the household but no reduction in household contacts [5]. The models predicted that although this partial shielding would be one of the most effective measures in reducing total deaths and severe outcomes, under the assumption of an infection fatality ratio of 0.9% it would not be enough to prevent critical care facilities from being overwhelmed.

A key objective of the emerging discipline of “precision medicine” is to use risk stratification to assist clinical decision making. This has driven forward methodological developments for instance in privacy-preserving analysis of information from medical records, and quantifying increments in predictive performance in ways that are relevant to decisions. As researchers in this field we have examined how risk stratification, based on using information from medical records to quantify risk of severe or fatal COVID-19, might lay the basis for a stratify-and-shield policy. We do not advocate a particular course of action but instead investigate the theoretical dependencies of the stratify-and-shield approach, its possible impact on mortality and workload for the health service, and discuss how to resolve some of the key uncertainties upon which the effectiveness of such an approach depends.

## Methods

### Modelling the stratify-and-shield policy with a probability tree

The stratify-and-shield policy can be modelled with a probability tree as shown in Figure 1. Those seronegative for auto-antibodies to to SARS-CoV-2 are classified as susceptible. Write *S* for the initial proportion of the population who are susceptible, and *F* for the infection fatality ratio (defined for this model as the proportion of susceptible individuals who will die if unshielded). Because information about the variation of infection fatality ratio by age and sex is accounted for in the classifier, we ignore it in the probability model and use only the population-wide infection fatality ratio. The classifier is applied to the susceptible population, considered as a mixture of “cases” (individuals who are destined to die if unshielded during the epidemic) and “non-cases” (individuals who are destined to survive if unshielded during the epidemic). Those at high risk are allocated to shielding until the epidemic among unshielded individuals is over, by which time the proportion of unshielded survivors who are immune has reached *I*_*u*_. We assume conservatively that by this time all unshielded individuals who were destined to die if unshielded have died.

**Fig 1.**
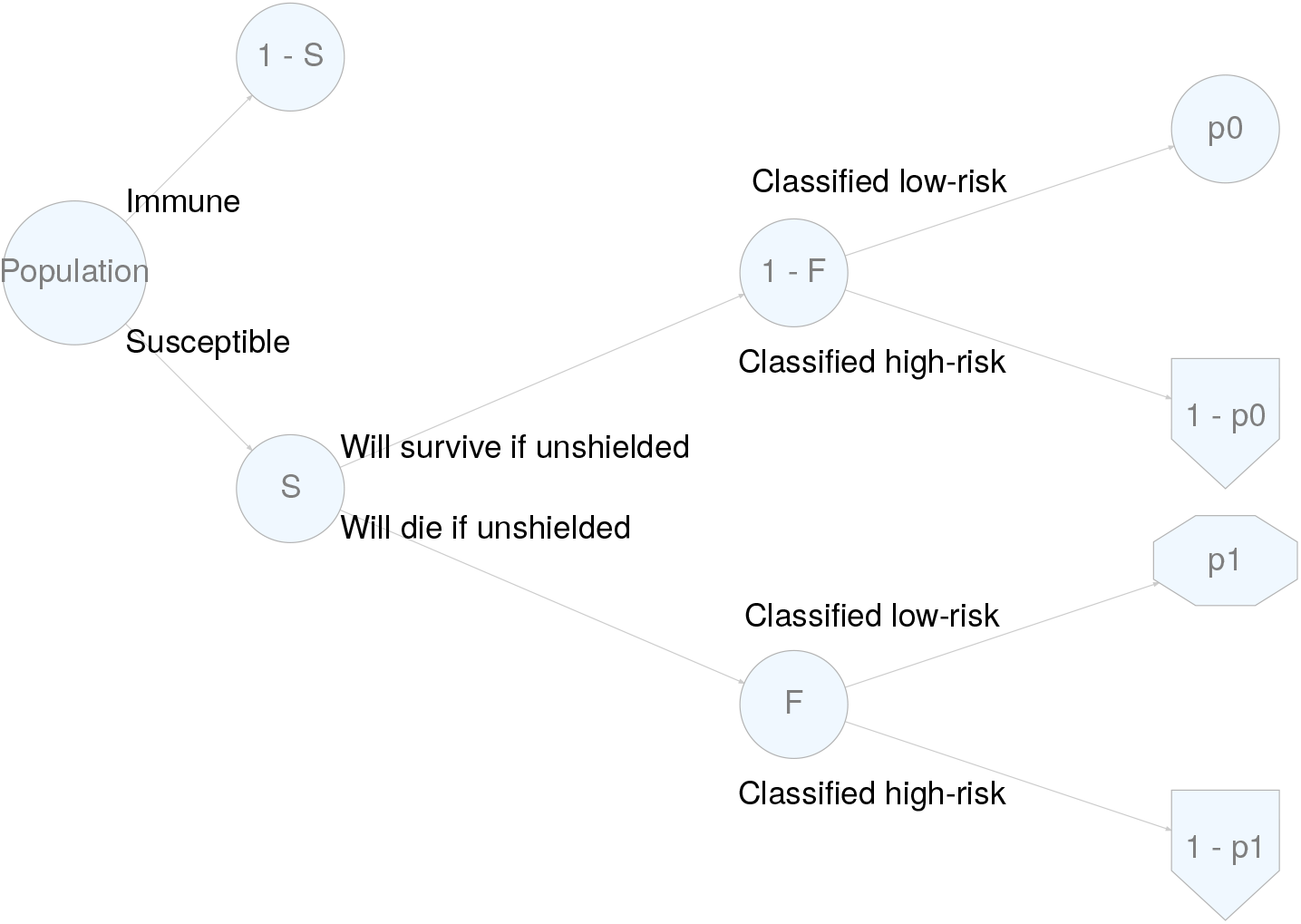
Probability tree for a stratify-and-shield policy

Write *p*_0_ for the proportion of non-cases correctly classified as low risk (specificity of the classifier) and *p*_1_ for the proportion of cases incorrectly classified as low risk (1 minus sensitivity of the classifier).

Of the original population:

- the proportion allocated to shielding is *S* [(1 *– F*) (1 *– p*_0_) + *F* (1 *– p*_1_)].
- the proportion initially susceptible and classified as low risk is *S* [(1 *– F*) *p*_0_ + *F*_*p*1_]
- the proportion who died because they were incorrectly classified as low risk and consequently unshielded is *SF*_*p*1_

The risk of death to susceptible individuals who are classified as low risk and consequently unshielded is

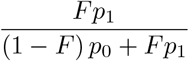

### Calculating the predictive performance of a risk classifier from the expected weight of evidence

To stratify people by risk, a classifier based on age, sex, other demographic variables and medical history can be constructed. For each individual, such a classifier will output the log-likelihood ratio (weight of evidence) favouring case over non-case status. To predict how such a classifier will behave when used for risk stratification, we can use the properties of the class-conditional sampling distributions of the weight of evidence favouring assignment to one class over the other. These distributions are constrained by mathematical consistency to have regularity properties [6]. The expectation of the weight of evidence favouring the correct over the incorrect class assignment is the expected information for discrimination conveyed by the classifier, denoted by the symbol Λ [7]. A key advantage of using Λ, rather than the familiar *C*-statistic (area under the receiver operating characteristic curve) to quantify classifier performance is that the contributions of independent predictors are additive on the scale of Λ. We can thus add the information for discrimination estimated from a case-control study matched on age and sex to the information for discrimination calculated from age- and sex-specific incidence rates.

If the classifier is constructed from many independent variables, the class-conditional sampling distributions of the weight of evidence favouring case over non-case status will be asymptotically Gaussian. In this situation the weight of evidence favouring case over non-case status in cases, and the weight of evidence favouring non-case over case status in non-cases will both be distributed with mean Λ and variance (when natural logarithms are used) 2Λ. As described elsewhere, this result allows us to calculate how the classifier will behave when used for risk stratification in the population, using only the value of Λ [7]. To facilitate intuitive interpretation, we use logarithms to base 2 so that Λ is expressed in bits (binary digits).

For a cut-off value of *w* bits for the weight of evidence favouring case over non-case status,

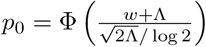 and 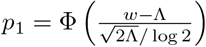, where Ф () is the cumulative distribution function of a standard normal variate.

For a given value of the classifier performance Λ, the infection fatality ratio *F*, and and the proportion *S* who are susceptible at the outset, we can find by iteration the value of *w* that will set the proportion of the population allocated to shielding to 0.15. From this we can calculate the specificity *p*_0_ and the sensitivity (1 – *p*_1_) of the classifier at this threshold.

### Threshold for population-level immunity and proportion of population to be shielded

The stratify-and-shield policy is intended to achieve population-level immunity quickly so that shielding of high-risk individuals can end. If in the surviving population at the end of the shielding period the proportion who were shielded is *P* and the proportion of unshielded individuals who are immune is *I*_*u*_*P*, the proportion *I*_*t*_ of the total population who are immune will be give by *I*_*t*_ = (1 – *P*) *I*_*u*_. The threshold value of *I*_*t*_ required to maintain population-level immunity is given by 1 – 1*/R*_0_, where *R*_0_ is the case reproduction number in a fully susceptible population [8]. This imposes a limit on the proportion of individuals who can be allocated to shielding, as otherwise when shielding ends the mixing of shielded individuals who are still susceptible will dilute the proportion *I*_*t*_ of the total population who are immune below this threshold.

Estimates of *R*_0_ have ranged from 2.5 [9] to 5.7 [10]. For this initial evaluation we assume *R*_0_ = 4, giving a threshold of *I*_*t*_ = 0.75, and we assume also that the final proportion *I*_*u*_ of unshielded individuals who are immune will be driven to 0.9 by overshoot of new infections after the threshold for population-level immunity has been reached. It follows that for *I*_*t*_ to be at least 0.75, *P* must be no more than 0.17. We have therefore set 15% as an upper limit for the proportion of the original population to be shielded under a stratify-and-shield policy.

### Relationship of the infection fatality ratio *F* to the proportion *S* who are susceptible at the start of the stratify-and-shield policy

Assuming that a single infection confers immunity in those who survive, the product of the infection fatality ratio *F* and the proportion 1 – *S* of individuals who are immune must equal the proportion of the total population who have died from the disease, allowing for lags from the date of infection to the date of death or development of immunity. In the UK by 18 April the proportion of the population that had died with confirmed COVID-19 was about 1 in 4500 individuals. For simplicity and to allow for overcounting of deaths in which COVID-19 was not the underlying cause, we assume that the proportion who have died from the disease at the start of shielding, equal to *F* (1 – *S*), is 1 in 5000. Thus varying the proportion (1 – *S*) of individuals initially immune from 20% to 5% corresponds to varying the infection fatality ratio *F* from 0.1% to 0.4%.

## Results

### Information for discrimination obtained with a classifier using age and sex only

We fitted a logistic regression model for the dependence of death from COVID-10 on age and sex, using the age-sex distribution of all deaths with mention of COVID-19 up to 18 April and population data for England and Wales. The odds ratio for death associated with a ten-year increase in age was 3 and the odds ratio associated with male sex was 2.1 The log-likelihood ratio favouring correct over incorrect assignment was computed by subtracting the log prior odds from the log posterior adds. Taking the average of this ratio (as the average of the mean in those who died and the mean in those who survived) gives an estimate for the expected information for discrimination of Λ = 3 bits. This provides a lower bound on a range of plausible values for the performance of a classifier that uses information from medical records in addition to age and sex: from associations with pre-existing morbidity we might expect to obtain between 0.5 and 1.5 bits of information for discrimination.

### Projected numbers of deaths in unshielded individuals

For each assumed pair of values for infection fatality ratio *F* and and initial proportion susceptible *S*, and each value of Λ, we calculated the threshold for sensitivity and specificity that would set the the proportion of the population allocated to shielding to 0.15. From this we calculate the expected number of deaths in unshielded individuals, under the conservative assumption that all those who are misclassified as low risk, and thus advised to forgo shielding, die before the epidemic ends. We also calculated the final proportion of the total population that is immune, given that the epidemic in the unshielded population continues until the fraction of the unshielded who remain susceptible at the end of the policy is 0.1 and that the proportion of the population allocated to shielding is 0.15 for reasons explained in the Method section. Readers can use the reader can use the code provided with this paper to evaluate other settings.

Table 1 shows the results of these calculations. The proportion *p*_1_ is the proportion of those destined to die who are incorrectly assigned to the unshielded group. The sensitivity (1 – *p*_1_) of the classifier, with threshold set so that 15% of the population will be classified as high risk, is the maximal proportion of deaths that can be prevented by a stratify-and shield-policy optimally applied, in comparison with an unselective lifting of social distancing. The results show that even under unfavourable assumptions – a classifier of modest performance and an infection fatality ratio as high as 0.4% – the sensitivity of the classifier is at least 75%.

**Table 1.** Mortality risk and expected deaths in the UK population of 66.65 million under an optimally applied stratify-and-shield policy, in which 15% of the population are shielded until 90% of unshielded individuals are immune

| Model assumptions |  |  | Model results |  |  |  |
| --- | --- | --- | --- | --- | --- | --- |
| Infection fatality risk $F$ | Initial proportion immune $(1 - S)$ | Expected information for discrimination $\Lambda$ (bits) | Specificity of classifier $p_0$ | Sensitivity of classifier $(1 - p_1)$ | Mortality risk in susceptible unshielded individuals | Expected deaths in unshielded individuals in UK population |
| 0.0010 | 0.20 | 3.0 | 0.81 | 0.79 | 1 / 5074 | 11130 |
| 0.0010 | 0.20 | 3.5 | 0.81 | 0.83 | 1 / 6160 | 9168 |
| 0.0010 | 0.20 | 4.0 | 0.81 | 0.86 | 1 / 7476 | 7553 |
| 0.0010 | 0.20 | 4.5 | 0.81 | 0.88 | 1 / 9075 | 6223 |
| 0.0015 | 0.13 | 3.0 | 0.83 | 0.77 | 1 / 2894 | 19514 |
| 0.0015 | 0.13 | 3.5 | 0.83 | 0.81 | 1 / 3492 | 16172 |
| 0.0015 | 0.13 | 4.0 | 0.83 | 0.84 | 1 / 4214 | 13402 |
| 0.0015 | 0.13 | 4.5 | 0.83 | 0.87 | 1 / 5086 | 11103 |
| 0.0020 | 0.10 | 3.0 | 0.83 | 0.77 | 1 / 2018 | 27987 |
| 0.0020 | 0.10 | 3.5 | 0.83 | 0.81 | 1 / 2428 | 23262 |
| 0.0020 | 0.10 | 4.0 | 0.83 | 0.84 | 1 / 2921 | 19331 |
| 0.0020 | 0.10 | 4.5 | 0.83 | 0.87 | 1 / 3517 | 16057 |
| 0.0040 | 0.05 | 3.0 | 0.84 | 0.75 | 1 / 907 | 62291 |
| 0.0040 | 0.05 | 3.5 | 0.84 | 0.79 | 1 / 1085 | 52023 |
| 0.0040 | 0.05 | 4.0 | 0.84 | 0.83 | 1 / 1300 | 43429 |
| 0.0040 | 0.05 | 4.5 | 0.84 | 0.86 | 1 / 1559 | 36233 |

If the population-wide infection fatality ratio is as low as 0.1%, then even with a classifier using that provides 3 bits of information for discrimination, equivalent to using only age and sex, the mortality risk in unshielded susceptible individuals is less than 1 in 5000, though the total expected deaths in unshielded individuals is about 11,000 in the UK population. Classifiers of higher performance reduce the risk to unshielded susceptible individuals and the total expected deaths in this group. Thus if the classifier provides 4 bits of information for discrimination, then for an infection fatality ratio of 0.1% the risk of death in unshielded individuals is about 1 in 7500 and the expected number of deaths in unshielded individuals is about 7600 applied to the UK population.

If however the infection fatality ratio is as high as 0.15%, then a classifier of higher performance – 4.5 bits – is required to keep the mortality risk in unshielded individuals below 1 in 5000. If the infection fatality ratio is 0.2% or more, it is not possible even using a classifier with Λ = 4.5 bits for a stratify-and-shield policy to keep the absolute risk in unshielded individuals below this level.

### Projected effect on critical care

From the ICNARC report on cases in critical care up to 16 April [11] we estimate that the fatality rate in those aged under 69 years admitted to critical care with COVID-19 is 44%. This implies that for every 10,000 deaths in the unshielded group, there will be about 24000 individuals requiring critical care. The average duration of stay of such cases is about one week in the ICNARC data. Additional cases would be expected in individuals offered shielding, either through failure of shielding or voluntary decision to forgo shielding. Health service managers have been set a target of expanding adult critical care capacity fourfold from a baseline of about 5000 beds before the coronavirus epidemic. If critical care capacity has been scaled up to provide 6000 additional beds, this should be able to deal with 24000 severe cases / month. The projections in Table 1 show that if the infection fatality ratio is no more than 0.2%, a classifier with Λ of at least 3.5 bits will keep the number of deaths in the unshielded group below about 25000 and the expected number requiring critical care below about 60,000.

### Comparison with adaptive social distancing

The adaptive social distancing policy is intended to limit the arrival rate of severe cases to a level that is within the capacity of critical care services to manage, and also to limit deaths by delaying infection until a vaccine is available [5]. As updates of the original modelling studies based on the rather lower values of the infection fatality ratio that are now considered plausible [12] have not been released, it is difficult to make direct comparisons between projections based on the adaptive social distancing policy and those based on the stratify-and-shield policy. The projections above suggest that unless the infection fatality ratio is higher than 0.2%, the stratify and shield policy should be able to limit the arrival rate of severe cases to a level that is within the capacity of a fourfold scale up of adult critical care facilities, if these arrivals are spread over about three months. The only situation, therefore, in which lockdowns lead to fewer deaths than the stratify-and-shield policy is if delaying the epidemic buys time for more effective treatments or vaccines to be developed.

An economic comparison is instructive. The Office of Budget Responsibility estimates that a three-month lockdown will reduce UK GDP by 35 percent for one quarter of the year [13]: this is about £194 billion. In the most favourable scenario for a three-month lockdown, the epidemic is suppressed to very low levels until an effective treatment or vaccine becomes available at the end of the lockdown. Under the least favourable scenario for the stratify-and-shield policy in Table 1, for an infection fatality ratio of 0.02 and a classifier based on age and sex only that extracts 3 bits of information the expected number of deaths in the unshielded group is 62291. This is the maximum number of deaths that could have been prevented by a lockdown in comparison to a stratify-and-shield policy. In this scenario, all males under age 60 and females under age 70 would have been allocated to the unshielded group. We can calculate an upper bound on the average years of life lost by those in this group who would have died as 27.3 years based on the (implausible) assumption that males under age 60 and females under age 70 who died with COVID-19 in England and Wales had the same life expectancy as those in the general population with the same age-sex distribution. We can use this to calculate a lower bound on the cost-effectiveness of the lockdown in comparison to stratify-and-shield as £114,000 per year of life gained under the most favourable scenario for the lockdown. For comparison, the National Institute of Clinical Excellence uses £30,000 per year of healthy life gained as a cutoff for deciding whether to recommend a new drug for the NHS.

## Discussion

We have shown that a stratify-and-shield policy would be constrained to shielding no more than 15% of the population, but that even if a classifier using medical history provided only a modest increment in predictive performance over one using only age and sex, it should be possible to allocate to the shielded group at least 80% of those who would die if unshielded. The average risk in those shielded, and the total number of deaths during the shielding period depend critically on the population-wide infection fatality ratio. If the infection fatality ratio is 0.4%, at the high end of the range of values we now consider plausible, even a relatively high-performing classifier would not be able to keep the number of deaths below 30,000 or the mortality risk in unshielded individuals below 1 in 2000 but this would still be within the capacity of an expanded critical care service to handle.

The level of risk that might be considered acceptable at a collective level in exchange for resuming economic and social activities is a policy decision. We note that the annual risk of accidental death in England and Wales for men and women aged 30-64 years is about 1 in 5000, and we do not stop the economy to prevent such deaths and neither do individuals stay home to avoid such a risk. We note also that the projected mortality risks in the unshielded are averages over the group; a risk classifier could of course output not simply a dichotomous classification, but a continuous risk score that would be the basis for individual choice. Any shielding strategy involves issues of ethics and equity in that those in the unshielded group are asked to accept a low risk so that not just they, but those shielded from infection, can emerge from isolation sooner.

It is to be expected that an epidemic in a susceptible population will overshoot the threshold for population-level immunity, as even when the reproduction number falls below 1 transmissions will continue for a few more generations till the epidemic is extinguished. This overshoot is usually considered undesirable [3], but for a stratify-and-shield policy it is desirable to drive the level of immunity as high as possible in the unshielded, so that when shielded individuals resume mixing the level of immunity in the total populatio is not diluted below the threshold for herd immunity.

This in turn implies that the epidemic in unshielded individuals should be allowed to run as fast as is compatible with not overloading critical care services, so as to maximize the overshoot. Keeping the duration of the shielding period short will also help to minimize the costs of shielding both to providers and to those shielded. The effect on the epidemic of lifting social distancing restrictions after a lockdown is hard to predict, and it is possible that the epidemic could peak rapidly such that half the severe cases expected in unshielded individuals arrive within one month. In this scenario, 20,000 is an upper limit on the total number of deaths during the shielding period that would not overload critical care services.

For projections of the impact of a stratify-and-shield policy, the key uncertainties are whether a single infection confers immunity, the infection fatality ratio, the predictive performance of the classifier, and the effectiveness of shielding over the few months required to allow the epidemic to run to completion in the low-risk group. We briefly consider what is the extent of these uncertainties and how they might be resolved.

### Does a single infection confer immunity?

Existing evidence on antibody-mediate immunity to coronaviruses has been reviewed elsewhere [14]. A study of seasonal coronavirus infections based on the Flu Watch cohort found no cases of reinfection with the same strain of coronavirus in a cohort of 216 adults with a first confirmed HCoV infection (0 of 8 individuals with two confirmed infections) [15]. In a study of 18 patients with SARS, IgG antibodies to SARS were reported to persist for at least 2 years [16]. Reinfection with SARS-CoV-2 could not be induced experimentally with macaquess [17]. Antibody-dependent enhancement of acute lung injury has been documented in experimental studies of SARS vaccines [18,19] but not with antibodies induced by infection with the SARS virus. As tens of thousands of people have now tested positive for SARS-CoV-2, the most direct way to establish whether repeat infection occurs is to link records of positive tests to identify recurrences after recovery from the first infection.

### Estimating the infection fatality ratio

The only direct estimate of the infection fatality ratio based on a population where all individuals were tested and followed for mortality is from the Diamond Princess cruise ship [20], where there were 7 deaths among 634 passengers who tested positive. Standardizing to the age structure of the England and Wales population would give a ratio of 0.1%. Indirect estimates are based on obtaining the numerator from reported deaths in a defined population, and the denominator from seroprevalence estimates. Stored samples from before December 2019 can be usd to control for the false positive rate of the antibody test. The Department of Health and Social Care reported on 4 April that Public Health England was “testing samples of the population and is in the process of analysing the first 800 samples collected” [21], but the results had not been published as of 25 April. Preliminary results of other seroprevalence surveys in Europe and the US have been reported. Though the reliability of some of these studies has been questioned [22] their findings are mostly consistent in showing seroprevalence levels around 3-4% in the first week of April 2020. As the time from SARS-CoV-2 exposure to seroconversion is estimated to be from 15 to 20 days [23], the proportion infected by late April is likely to be considerably higher than the seroprevalence in early April.

### Practical feasibility of a stratify-and-shield policy

A detailed examination of the practical feasibility of a stratify-and-shield policy is beyond the scope of this paper. Compliance (assumed to be voluntary) with shielding in the high-risk group must be high enough that severe cases among those who forgo shielding do not overload the health service, but unlike the adaptive social distancing policy the stratify-and-shield policy would not require coercion. It does however require that support for shielding is adequate to protect high-risk individuals from infection while the epidemic in the unshielded group continues; if this support were implemented in a half-hearted manner, the outcome for those who rely on social care would be poor. Intense shielding is already recommended in UK guidance for more than 1 million people who are “clinically extremely vulnerable” because they have received organ transplants, are on immunosuppressants or chemotherapy for cancer, have severe respiratory conditions, or have rare diseases that increase the risk of infections [24]. These individuals are advised to adopt rigorous shielding measures, including staying at home at all times, isolating themselves from others in the same household and not going out even for food or medicine. Assistance with delivery of food and medicines is offered to people in this group.

The logistic challenges of providing this level of shielding to 15% of the population are formidable, but not necessarily insurmountable if the duration of shielding is only a few months. For instance, high-risk adults in crowded households might have to be offered alternative accommodation until low-risk household members are immune, and paid caregivers who are not already immune would need to be trained and equipped to use reverse barrier precautions. Serological testing could be used to identify individuals who are no longer susceptible as “focal points for sustaining safer interactions” with those who are shielded [25]. Shielding of individuals who are in employment would require guarantees that their employment status would not be jeopardized by isolating themselves for a few months.

## Conclusion

A stratify-and-shield policy using a classifier based on medical records has the potential to save lives, restore economic activity and end the epidemic long before a vaccine is expected to be available. The key uncertainties about the theoretical impact of this policy – the infection fatality ratio, the extent to which infection confers immunity, and the performance of a classifier – should be resolved by studies now under way. This policy option should not be dismissed but seriously evaluated as an alternative to adaptive social distancing.

## Data Availability

All source code is available form the authors on request. Only publicly available data are used.

## Notes

### Competing Interest Statement

The authors have declared no competing interest.

### Funding Statement

none

## References

1. Scientific Advisory Group for Emergencies. SPI-M-O: Consensus view on behavioural and social interventions. https://assets.publishing.service.gov.uk/government/up-loads/system/uploads/attachment_data/file/873729/06-spi-m-o-consensus-view-on-behavioural-and-social-interventions.pdf; 2020 Mar.

2. Peto J, Alwan NA, Godfrey KM, Burgess RA, Hunter DJ, Riboli E, et al. Universal weekly testing as the UK COVID-19 lockdown exit strategy. The Lancet. 2020;0. doi:10.1016/S0140-6736(20)30936-3

3. Handel A, Miller J, Ge Y, Fung IC-H. If long-term suppression is not possible, how do we minimize mortality for COVID-19 and other emerging infectious disease outbreaks? medRxiv. 2020; 2020.03.13.20034892. doi:10.1101/2020.03.13.20034892

4. Scientific Advisory Group for Emergencies. Potential effect of non-pharmaceutical interventions (NPIs) on a COVID-19 epidemic in the UK. https://assets.publishing.service.gov.uk/government/uploads/system/uploads/attach-ment_data/file/873723/03-potential-effect-of-non-pharmaceutical-interventions-npis-on-a-Covid-19-epidemic-in-the-UK.pdf; 2020 Feb.

5. Ferguson NM, Nedjati-Gilani G, Imai N, Ainslie K, Baguelin M. Report 9: Impact of non-pharmaceutical interventions (NPIs) to reduce COVID-19 mortality and healthcare demand. https://www.imperial.ac.uk/media/imperial-college/medicine/mrc-gida/2020-03-16-COVID19-Report-9.pdf: Imperial College COVID-19 Response Team; 2020 Mar.

6. Good IJ, Toulmin GH. Coding theorems and weight of evidence. Journal of the Institute of Mathematics and Applications. 1968;4: 94–105.

7. McKeigue P. Quantifying performance of a diagnostic test as the expected information for discrimination: Relation to the C-statistic. Stat Methods Med Res. 2019;28: 1841–1851. doi:10.1177/0962280218776989

8. Fine P, Eames K, Heymann DL. ”Herd immunity”: A rough guide. Clin Infect Dis. 2011;52: 911–916. doi:10.1093/cid/cir007

9. Anderson RM, Heesterbeek H, Klinkenberg D, Hollingsworth TD. How will country-based mitigation measures influence the course of the COVID-19 epidemic? Lancet. 2020;395: 931–934. doi:10.1016/S0140-6736(20)30567-5

10. Sanche S, Lin YT, Xu C, Romero-Severson E, Hengartner N, Ke R. Early Release - High Contagiousness and Rapid Spread of Severe Acute Respiratory Syndrome Coronavirus 2 - Volume 26, Number 7July 2020 - Emerging Infectious Diseases journal - CDC. 2020. doi:10.3201/eid2607.200282

11. ICNARC. ICNARC report on COVID-19 in critical care. https://assets.publishing.service.gov.uk/government/uploads/system/uploads/attach-ment_data/file/873729/06-spi-m-o-consensus-view-on-behavioural-and-social-interventions.pdf: Intensive Care National Audit and Research Centre; 2020 Apr.

12. Oke J, Heneghan C. Global Covid-19 Case Fatality Rates. Centre for Evidence-Based Medicine, Oxford UniversityBM. https://www.cebm.net/covid-19/global-covid-19-case-fatality-rates/; 2020.

13. Office for Budget Responsibility. Commentary on the OBR coronavirus reference scenario. https://obr.uk/download/coronavirus-reference-scenario-the-obrs-coronavirus-analysis/; 2020 Apr.

14. Huang AT, Garcia-Carreras B, Hitchings MDT, Yang B, Katzelnick L, Rattigan SM, et al. A systematic review of antibody mediated immunity to coronaviruses: Antibody kinetics, correlates of protection, and association of antibody responses with severity of disease. medRxiv. 2020; 2020.04.14.20065771. doi:10.1101/2020.04.14.20065771

15. Aldridge RW, Lewer D, Beale S, Johnson AM, Zambon M, Hayward AC, et al. Seasonality and immunity to laboratory-confirmed seasonal coronaviruses (HCoV-NL63, HCoV-OC43, and HCoV-229E): Results from the Flu Watch cohort study. Wellcome Open Res. 2020;5: 52. doi:10.12688/wellcomeopenres.15812.1

16. Mo H, Zeng G, Ren X, Li H, Ke C, Tan Y, et al. Longitudinal profile of antibodies against SARS-coronavirus in SARS patients and their clinical significance. Respirology. 2006;11: 49–53. doi:10.1111/j.1440-1843.2006.00783.x

17. Bao L, Deng W, Gao H, Xiao C, Liu J, Xue J, et al. Reinfection could not occur in SARS-CoV-2 infected rhesus macaques. bioRxiv. 2020; 2020.03.13.990226. doi:10.1101/2020.03.13.990226

18. Tseng C-T, Sbrana E, Iwata-Yoshikawa N, Newman PC, Garron T, Atmar RL, et al. Immunization with SARS coronavirus vaccines leads to pulmonary immunopathology on challenge with the SARS virus. PLoS ONE. 2012;7: e35421. doi:10.1371/journal.pone.0035421

19. Liu L, Wei Q, Lin Q, Fang J, Wang H, Kwok H, et al. Anti-spike IgG causes severe acute lung injury by skewing macrophage responses during acute SARS-CoV infection. JCI Insight. 2019;4. doi:10.1172/jci.insight.123158

20. Russell TW, Hellewell J, Jarvis CI, van Zandvoort K, Abbott S, Ratnayake R, et al. Estimating the infection and case fatality ratio for coronavirus disease (COVID-19) using age-adjusted data from the outbreak on the Diamond Princess cruise ship, February 2020. Euro Surveill. 2020;25. doi:10.2807/1560-7917.ES.2020.25.12.2000256

21. Department of Health and Social Care. Coronavirus (COVID-19) Scaling up our testing programmes. https://assets.publishing.service.gov.uk/government/uploads/sys-tem/uploads/attachment_data/file/878121/coronavirus-covid-19-testing-strategy.pdf; 2020 Apr.

22. Vogel G. Antibody surveys suggesting vast undercount of coronavirus infections may be unreliable. Science | AAAS. 2020;doi:10.1126/science.abc3831.

23. Lou B, Li T, Zheng S, Su Y, Li Z, Liu W, et al. Serology characteristics of SARS-CoV-2 infection since the exposure and post symptoms onset. medRxiv. 2020; 2020.03.23.20041707. doi:10.1101/2020.03.23.20041707

24. Public Health England. Guidance on shielding and protecting people who are clinically extremely vulnerable from COVID-19. https://obr.uk/download/coronavirus-reference-scenario-the-obrs-coronavirus-analysis/; 2020.

25. Weitz JS, Beckett SJ, Coenen AR, Demory D, Dominguez-Mirazo M, Dushoff J, et al. Intervention Serology and Interaction Substitution: Modeling the Role of ‘Shield Immunity’ in Reducing COVID-19 Epidemic Spread. medRxiv. 2020; 2020.04.01.20049767. doi:10.1101/2020.04.01.20049767

